# Population-level trends of Psychiatric Medication Co-Prescriptions in Persons with Epilepsy: an EPIC Cosmos Study

**DOI:** 10.64898/2026.07.24.26358856

**Authors:** Hannah Kostan, Vaishnav Krishnan

## Abstract

Persons with epilepsy (PWE) experience high rates of psychiatric comorbidity, yet the population-scale pharmacoepidemiology of psychiatric medication (PM) use alongside antiseizure medications (ASMs) has not been previously characterized. Using Epic Cosmos, a federated electronic health record network spanning >300 million patients across >2,000 hospital systems, we examined patterns of ASM and PM co-prescriptions in PWE (ICD-10 G40.x) every year between 2018–2025. PM prescriptions were similarly assessed in patients with asthma (J45.x). We found that despite the introduction of several newer ASMs, the overall prescribing landscape remained stable, with little change in the relative use of individual ASMs over time. Compared with asthma patients, PWE were more likely to receive prescriptions for opiates, antidepressant and antipsychotic medications across the age spectrum. ≤17-year-old PWE were more likely to receive ADHD/stimulant medications, whereas adults and older adults exhibited a shift toward cognitive enhancing agents. We did not observe a preponderance of PM co-prescribing with any specific ASM or ASM class. Together, these results provide a population-scale, age-specific survey of psychiatric medication co-prescriptions in epilepsy, establishing a framework to monitor surrogate markers of psychiatric comorbidity and to support pharmacovigilance of potential drug-drug interactions.

## Introduction

Psychiatric comorbidity is among the most consequential and undertreated dimensions of epilepsy^1^. Conditions such as depression, anxiety, psychosis, and attention-deficit/hyperactivity disorder occur in persons with epilepsy (PWE) at rates substantially exceeding those of the general population, and independently predict poorer seizure control, psychiatric side effects to antiseizure medications (ASMs) and reduced quality of life^2-5^. Whether they arise before or after seizure onset, managing these comorbidities frequently requires co-prescriptions for psychiatric medications (PMs). In PWE who are already prescribed one or more ASMs, PMs augment the problem of CNS polytherapy and can lead to poorer treatment adherence and greater overall costs^6,7^. Some PMs can lower seizure threshold (e.g., bupropion), and many display clinically meaningful pharmacokinetic interactions with enzyme-inducing or enzyme-inhibiting ASMs^8,9^. Thus far, assessments of PM co-prescribing in PWE have derived from single-center adult or pediatric cohorts of limited size^10,11^, precluding broader and more universal insights.

Here, we characterize patterns of ASM and PM co-prescribing at a previously unachievable scale by leveraging Epic Cosmos^12^, a federated electronic health record (EHR) research platform that aggregates de-identified data belonging to more than 300 million patients from over ∼2000 hospital systems based predominantly in the US. Over a 7-year study period (2018-2025) and across three major age groups (≤17, 18-64 and ≥65-year-olds), we (i) examine trends in the overall landscape of ASM prescriptions, (ii) illustrate how PM co-prescribing rates are amplified in PWE compared with those diagnosed with asthma, and (iii) explore whether specific classes of PMs are co-prescribed with specific ASMs across the age spectrum.

## Methods

In March 2026, we queried Epic Cosmos, a HIPAA-defined dataset derived from the EHRs of participating Epic health systems. Cosmos supports cohort definitions by diagnosis code, demographic stratification, and medication-exposure counts. Because our analyses used aggregate, de-identified counts, this work was exempt from institutional review board oversight. Queries were performed separately for each of 8 calendar years (2017-2025) and stratified into three age bands (≤17, 18–64, and ≥65 years). For each year, persons with epilepsy (PWE) were defined as those assigned an ICD-10 diagnosis of epilepsy (G40.x) for an admission, billing or encounter diagnosis, *and* who were dispensed a prescription for at least one of 31 ASMs (including benzodiazepines). We excluded epilepsy diagnoses reported on “Problem Lists”, which can be biased by the inaccuracies of self-reporting. We similarly assembled annual cohorts of patients with asthma (J45.x), selected as a chronic, highly prevalent non-neurological condition managed within similar inpatient and ambulatory care settings. Asthma has also been historically employed to illustrate the higher incidence of psychiatric comorbidities in PWE^13,14^. For both cohorts, we catalogued the total number of prescriptions for each of 54 PMs that belonged to six broad pharmacological classes: opiates, sleep aids, stimulants/ADHD treatments, antipsychotics, antidepressants/anxiolytics, and cognitive enhancers. We used the proportion of patients (*p*) prescribed each PM to compute odds (*p*/1-*p*) and odds ratios (comparing patients with epilepsy to those with asthma). Statistically significant differences in prescribing proportions were evaluated using two-proportion Z-tests^15^, with Bonferroni-adjusted p-values used to account for multiple comparisons (statistical significance was defined as p<0.05). Within PWE, we further derived the proportion of patients receiving each PM for each ASM assessed every year (data from 2025 is shown). Queries resulting in a small number of counts (e.g., ≥65-year-olds prescribed fenfluramine) were censored as “fewer than 10 subjects” per Cosmos reporting policy.

## Results

The total number of PWE captured annually in Epic Cosmos, defined by the presence of both a qualifying ICD diagnosis *and* a dispensed ASM prescription increased from ∼900,000 to ∼1.6 million between 2018 and 2025 (Fig. 1A). 18-64-year-olds consistently comprised of ∼60% of the cohort. The total number of patients with asthma also expanded, from ∼4.4 to ∼8.4 million patients over the same study period. Rather than indicating a true increase in disease prevalence, these findings more likely reflect the dynamic nature of Epic Cosmos, with participating health systems continually contributing and updating data^16^. Within PWE, we examined the relative frequency of prescriptions for 31 ASMs (including benzodiazepines). As shown in Fig. 1B, levetiracetam had the highest share of ASMs across all age groups and years studied, amounting to ∼22% of all ASM prescriptions in ≤ 64-year-olds, and ∼33% of ASMs in ≥65-year-olds. Despite the advent of several new ASMs, including cenobamate, cannabidiol, fenfluramine, ganaxolone and stiripentol, the overall landscape of ASM prescriptions remained largely stable.

**Figure 1.**
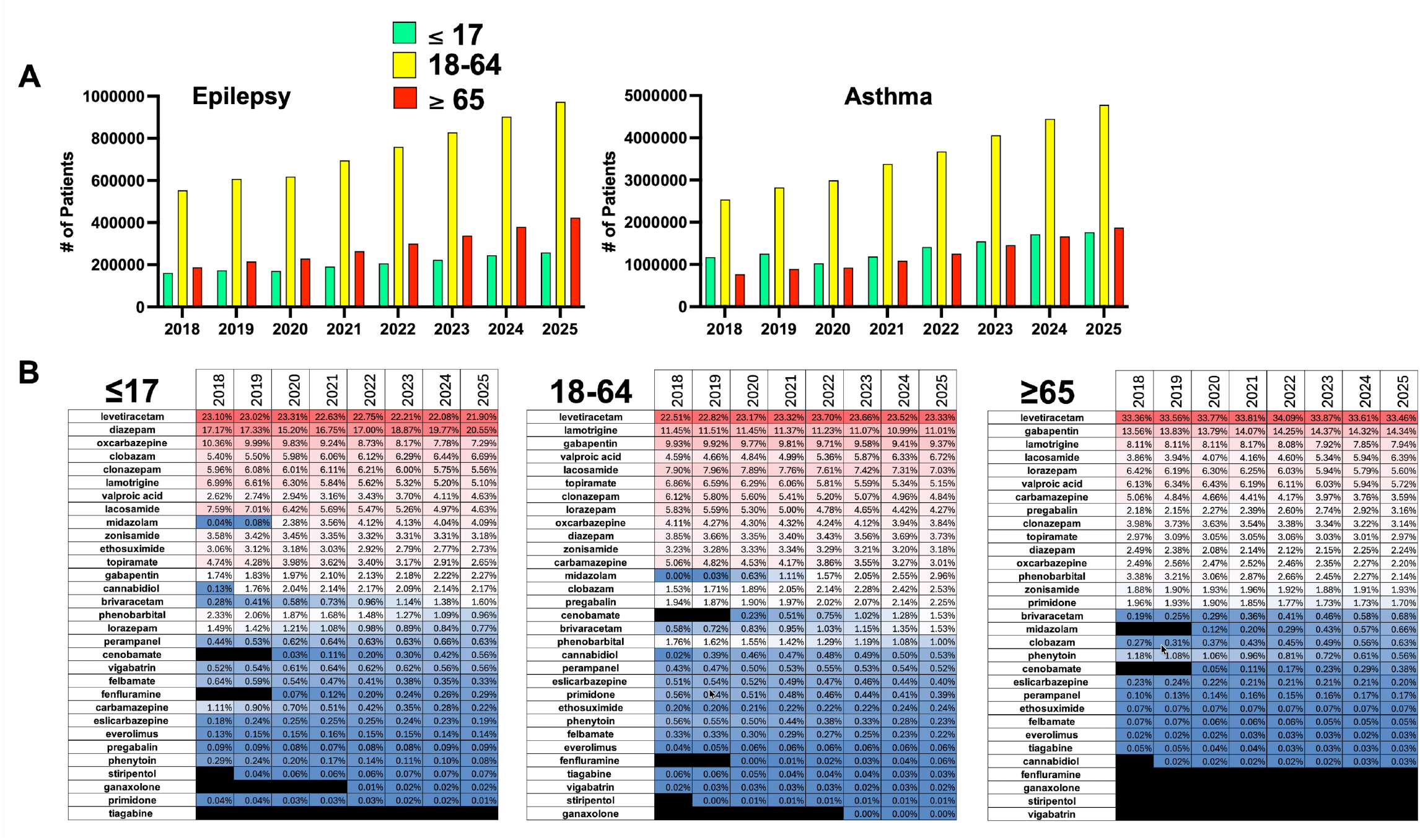
Epilepsy Patients and ASMs Prescribed. (A) Annual numbers of patients meeting study definitions for epilepsy (left) and asthma (right) shown, stratified by age (note that y-axes are not identical). (B) Proportion of all ASM prescriptions for PWE, grouped by year and age group. Within each table, ASMs are ranked by decreasing frequency of use as measured in 2025.

Figure 2 depicts a heatmap of odds ratios designed to illustrate the relative enrichment (or underutilization) of specific PMs in patients with epilepsy over asthma, assessed annually between 2017 and 2025. Across the age spectrum, PWE were more likely to receive PM prescriptions across a wide range of medication classes. Within ≤17-year-olds, we observed a marked increase in the use of memantine, frequently used as an off-label treatment for autism spectrum disorder^17^. Compared with older groups, ≤17-year-old patients featured greater ORs for the use of sleep aids, stimulants and a wide range of antipsychotic medications. Opiates were prescribed ∼1.3–1.7-fold more often in epilepsy among adults. By contrast, most antidepressants/anxiolytics drugs showed only modest relative increases in PWE (mean OR ≈ 1.1–1.2 in adults), with the exception of mirtazapine (OR ≈ 2). Compared with ≤17-year-olds, “Z-drug” sleep aids (zaleplon, eszopiclone, zolpidem) were significantly less likely to be prescribed older PWE. Bupropion, a norepinephrine/dopamine reuptake inhibitor known to lower seizure threshold, was appropriately under prescribed to PWE across all age groups and years studied.

**Figure 2.**
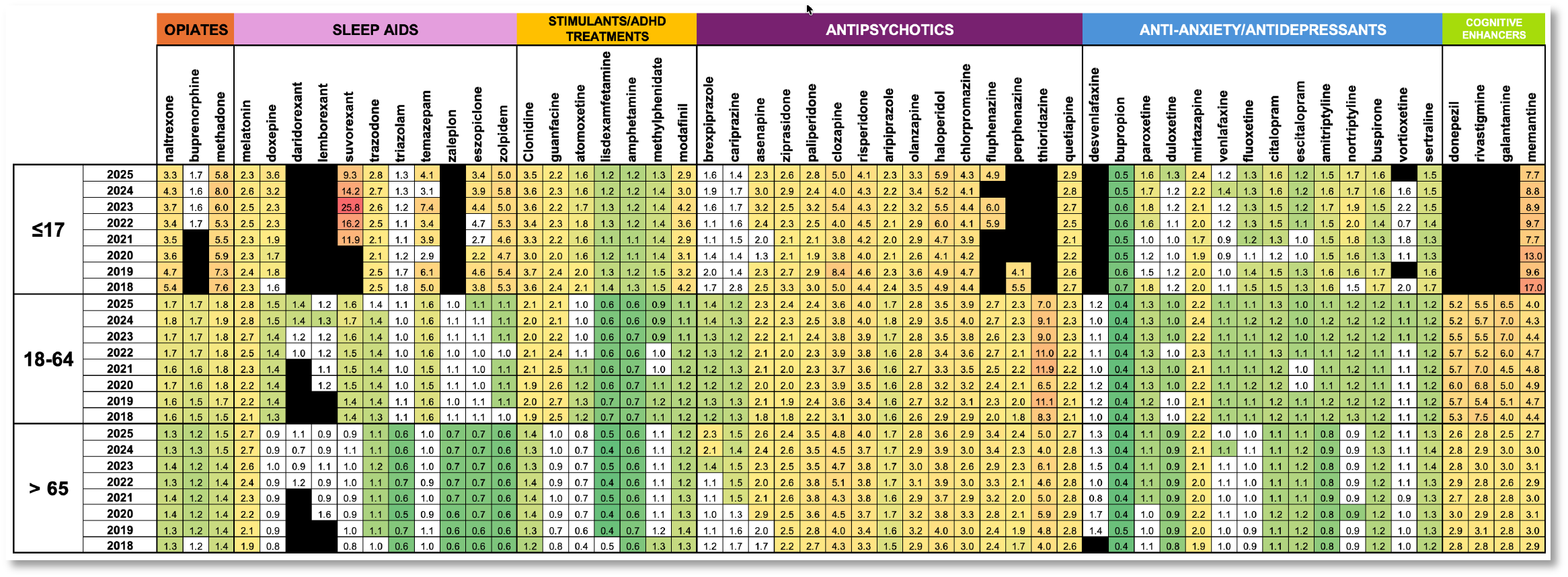
Odds Ratios (ORs) for PM prescriptions in PWE compared with asthma. For the population of PWE depicted in Fig. 1A, this heatmap tabulates the ORs of PM co-prescription in PWE over asthma. Colored cells reflect statistically significant changes (two proportion Z-tests, Bonferroni corrected for multiple comparisons). Blacked out cells reflect censored data, related to too few prescriptions in either PWE or asthma cohorts.

Next, we asked whether PM prescription rates varied by ASM (or ASM class). Figure 3 combines data from ∼ 1.5 million ASM and another ∼1.5 million PM prescriptions in ∼1.6 million PWE in 2025 (Fig. 1). ≤17-year-olds received high rates of prescriptions for ADHD medications, with clonidine being the most frequently prescribed agent. In comparison, antidepressants/anxiolytic agents were more heavily utilized in 18-64 and ≥65 age groups, with sertraline being the most frequently prescribed agent. Among patients ≥65 of age, prescriptions for cognitive enhancers became increasingly common, consistent with the greater prevalence of dementia in older adults with epilepsy. Across all age groups, trazodone emerged as the most commonly co-prescribed sleep aid. Despite the known negative psychotropic effects of levetiracetam^3,10^, PM prescription rates were not consistently higher in this group compared with other ASM classes, including sodium channel inhibitors. Overall, these data revealed that age (rather than ASM or ASM class) was the strongest determinant of PM co-prescribing.

**Figure 3.**
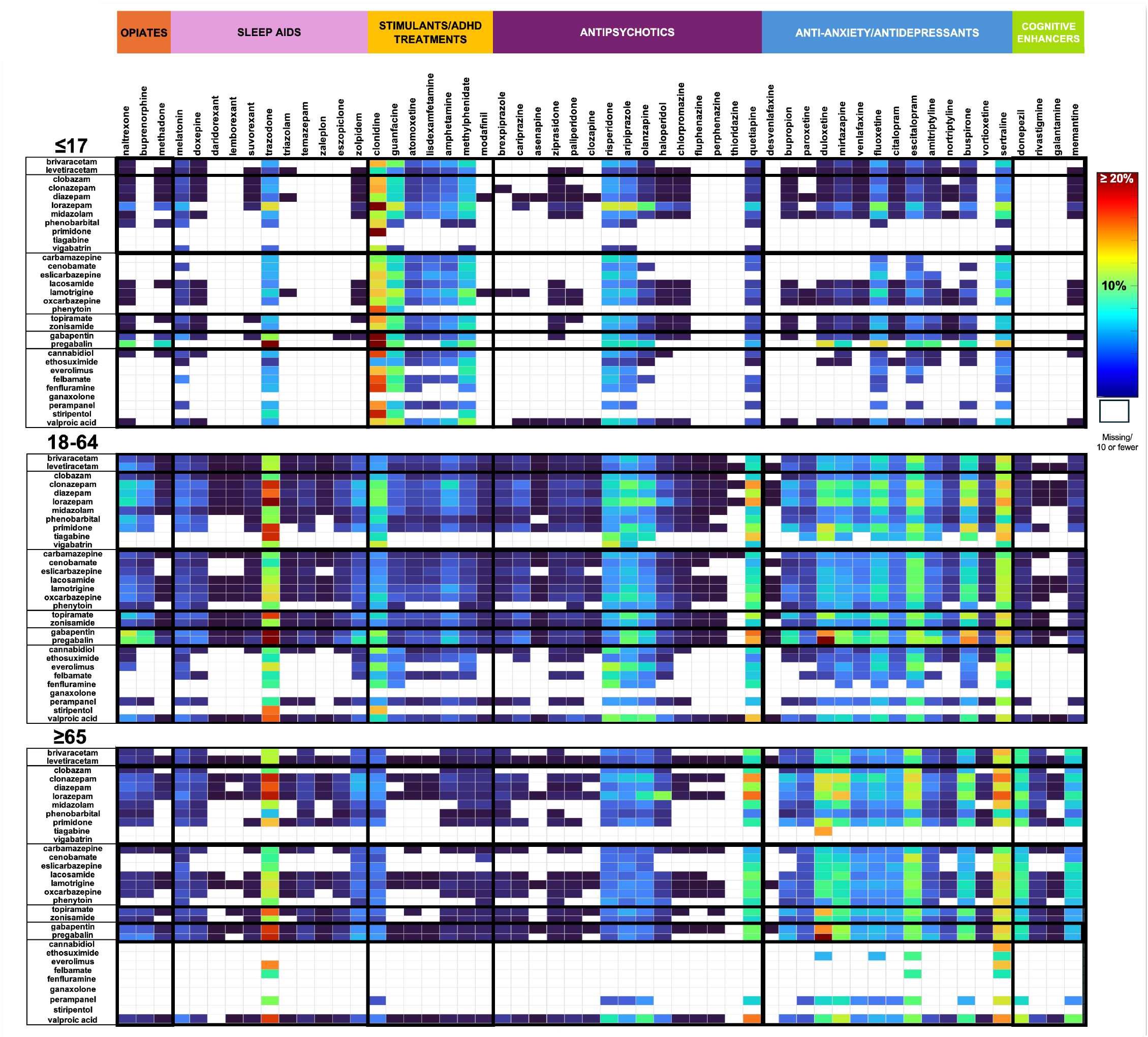
PM Co-prescriptions by Age and ASM/ASM class. Heatmap reflecting the color-coded percentage of PWE receiving a specific ASM (x-axis) that also received a co-prescription for a specific PM (y-axis) in 2025. White cells are censored data relating to too few prescriptions for either ASM or PM. We defined six major classes, including SV2a antagonists (levetiracetam, brivaracetam), GABAergic agents (e.g., clobazam, clonazepam), sodium channel blockers (e.g., phenytoin, carbamazepine), carbonic anhydrase inhibitors (topiramate/zonisamide), calcium channel blockers (gabapentin/pregabalin) and a final “other” class containing drugs that employ a mixture of other mechanisms.

## 4. Discussion

In this study, we leveraged a federated EHR network (Epic Cosmos) to explore recent trends in ASM and PM co-prescribing patterns in PWE at an unprecedented scale. We report three main findings. First, by surveying the proportion of annually dispensed ASM prescriptions between 2018-2025, we reveal a relative inertia in ASM prescribing landscapes. Levetiracetam was the most prescribed ASM across all age groups, accounting for approximately 1 in 5 prescriptions among patients ≤64 years old and 1 in 3 prescriptions among those ≥65 years old. The composition of the top four to five prescribed ASMs remained largely stable across age groups during the study period. Second, compared to patients with asthma, the relative increases (or decreases) in PM co-prescriptions in PWE varied markedly by age. In addition to stimulants/ADHD treatments and sleep aids, ≤17-year-olds featured higher odds of prescriptions for naltrexone, methadone and memantine, which may point to the heightened co-occurrence of conditions such as opiate use disorder, certain eating disorders and autistic/self-injurious behaviors. In adult patients, a relative decline in ADHD medication co-prescribing was accompanied by the increased prescribing of cognitive enhancers, which were prescribed approximately 4–5 times more frequently among 18–64-year-olds and approximately 3 times more frequently in ≥65-year-olds. These findings reflect the greater use of medications designed to address memory and cognitive symptoms in aging PWE. In contrast, across PWE from all age groups, we observed a relative enrichment in prescriptions for melatonin (∼2 fold) and antipsychotics (∼2-3 fold), as well as significantly fewer prescriptions for bupropion (∼2-fold reduction). Third, we did not observe a relative enrichment of PM co-prescribing with any specific ASM or ASM class. Together, our results provide insights into the burden of psychiatric comorbidity that are orthogonal to approaches that purely rely on ICD codes for case ascertainment, especially since the initiation of treatment for psychiatric illness maybe a surrogate marker of greater disease severity^1,18^.

The principal strength of this work is statistical power, providing access to a large, diverse multi-institutional patient population^19^. By drawing from hundreds of thousands of patients, Epic Cosmos can reveal overarching prescribing practices at a scale that is unattainable in single-center or regional studies, doing so across age strata and successive years. This scale is particularly valuable for visualizing patterns in pharmacologically important but new or uncommonly prescribed agents (e.g., everolimus or suvorexant). Our study is also limited by several factors. Our cohorts are exclusively derived from health systems that employ Epic and elect to participate in Cosmos^20^. Our reliance on ICD10 coding may inadvertently include patients with seizure mimics, such as functional seizures, where PM co-prescriptions are also common. Cosmos captures dispensed prescriptions but not adherence, and our analysis does not explore patterns of ASM polytherapy. We also cannot reliably discern which conditions or symptom clusters were addressed with specific PMs. Finally, aggregate counts preclude patient-level adjustment for confounders such as socioeconomic status, epilepsy etiology, or seizure severity.

Overall, this study provides a contemporary, population-scale view of ASM and PM co-prescribing in PWE, revealing stable ASM prescribing practices alongside substantial age-dependent differences in psychiatric medication use. These findings establish a benchmark for future studies of treatment patterns and highlight the potential of large federated EHR networks to generate clinically meaningful insights into the evolving pharmacologic management of epilepsy and its psychiatric comorbidities.

## Data Availability

All data produced in the present study are available upon reasonable request to the authors, and can be independently obtained through Epic Cosmos.

